# Physical activity, BMI and COVID-19: an observational and Mendelian randomisation study

**DOI:** 10.1101/2020.08.01.20166405

**Authors:** Xiaomeng Zhang, Xue Li, Ziwen Sun, Yazhou He, Wei Xu, Harry Campbell, Malcolm G Dunlop, Maria Timofeeva, Evropi Theodoratou

**Author notes:** Corresponding authors Evropi Theodoratou, Centre for Global Health, Usher Institute, University of Edinburgh, Edinburgh, United Kingdom, Maria Timofeeva, Colon Cancer Genetics Group, Institute of Genetics and Molecular Medicine, University of Edinburgh, Edinburgh, United Kingdom.

## Abstract

Physical activity (PA) is known to be a protective lifestyle factor against several non-communicable diseases while its impact on infectious diseases, including Coronavirus Disease 2019 (COVID-19) is not as clear. We performed univariate and multivariate logistic regression to identify associations between body mass index (BMI) and both objectively and subjectively measured PA collected prospectively and COVID-19 related outcomes (Overall COVID-19, inpatient COVID-19, outpatient COVID-19, and COVID-19 death) in the UK Biobank (UKBB) cohort. Subsequently, we tested causality by using two-sample Mendelian randomisation (MR) analysis. In the multivariable model, the increased acceleration vector magnitude PA (AMPA) was associated with a decreased probability of overall and outpatient COVID-19. No association was found between self-reported moderate-to-vigorous PA (MVPA) or BMI and COVID-19 related outcomes. Although no causal association was found by MR analyses, this may be due to limited power and we conclude policies to encourage and facilitate exercise at a population level during the pandemic should be considered.

## Introduction

Coronavirus Disease 2019 (COVID-19) caused by severe acute respiratory syndrome coronavirus 2 (SARS-CoV-2) was declared a pandemic on Mar 11^th^, 2020 by the World Health Organisation (WHO). By July 29^th^, 2020, a total of 16,558,289 patients had been diagnosed, with 656,093 confirmed deaths globally.^1^ National responses to the pandemic including a range of non-pharmacological interventions [NPIs], have been adopted by countries at nation-wide, state-wide, or city-wide levels. The restrictions imposed by governments during the outbreak have had a substantial impact on patterns of physical activity (PA) within populations. For example, an online survey conducted in Canada indicated that 40.5% of physically inactive individuals became less active and 33% became more active; 22.4% of physically active individuals became less active and 40.3% became more active during lockdown time.^2^

PA is known to be a protective lifestyle factor for a number of non-communicable diseases (e.g. cancer and cardiovascular disease) and aging processes (e.g. immunosenescence).^3-5^ However, evidence of the role of PA on respiratory viral infections remains weak, especially for highly contagious viruses like SARS-CoV-2. Since doing exercise could compromise social distancing measures and increase opportunities for contracting the virus (probably indoors more than outdoors), it is unclear whether being physically active is a beneficial lifestyle factor for respiratory viral infections. Viral pathogenesis has been reported to be greater in obese or overweight individuals^6^ and obesity has been suggested as a mediator for PA effects. In this study, we analysed whether PA and body mass index (BMI) influence the risk of COVID-19 and then investigated any causal associations between PA, BMI, and COVID-19 by applying two-sample Mendelian Randomisation (MR) analysis.

## Results

The characteristics of participants in the four outcome groups (overall COVID-19, inpatient COVID-19, outpatient COVID-19, and COVID-19 death), and the included controls were described in Table 1. Patients who died from COVID-19 had the highest average age (74.66) while outpatients had the lowest average age (65.97). The proportion of males was the highest for the COVID-19 death group (63.91%) and the lowest for the control group (45.57%). Participants in the inpatient COVID-19 group had the longest MVPA time while participants in the outpatient group had the shortest. For AMPA, participants in the control group have the longest AMPA time while patients who died from COVID-19 have the shortest. The results of univariate and multivariate logistic regression of two types of PA on the four outcomes are presented in Table 2. The results of univariate and multivariate logistic regression of BMI on the four outcomes are presented in Table S1. The results of two-sample MR analyses are presented in Figure 1 and Table S2.

**Table 1.** Characteristics of UK Biobank participants.

| Table 1 Characteristics of UK Biobank participants |  |  |  |  |  |  |
| --- | --- | --- | --- | --- | --- | --- |
|  | Overall COVID-19 cases<br>N (%) / Mean (SD) | Inpatient COVID-19 cases<br>N (%) / Mean (SD) | Outpatient COVID-19 cases<br>N (%) / Mean (SD) | COVID-19 deaths<br>N (%) / Mean (SD) | Controls<br>N (%) / Mean (SD) | Controls*<br>N (%) / Mean (SD) |
| All participants | 1746 (0.35%) | 1020 (0.20%) | 576 (0.11%) | 399 (0.07%) | 500758 (99.65%) | 415596 (99.65%) |
| Age | 68.76 (9.18) | 69.40 (8.86) | 65.97 (9.41) | 74.66 (5.98) | 68.46 (8.11) | 68.14 (8.08) |
| Sex | Male (52.92%) | Male (55.88%) | Male (45.83%) | Male (63.91%) | Male (45.57%) | Male (44.63%) |
| MVPA (MET-minutes/week) | 990.40 (1310.75) | 1039.00 (1356.07) | 898.7 (1236.67) | 1017.00 (1057.92) | 973.80 (1269.00) | 974.60 (1268.05) |
| AMPA (milli-gravities) | 26.72 (8.57)<br>(n=215) | 26.59 (8.85)<br>(n=122) | 27.38 (8.08)<br>(n=79) | 24.13 (8.10)<br>(n=36) | 28.00 (8.24)<br>(n=96460) | 28.12 (8.19)<br>(n=83748) |
| BMI (kg/m<sup>2</sup>) | 27.53 (4.88) | 27.47 (4.81) | 27.66 (5.01) | 27.59 (4.68) | 27.41 (4.79) | 27.41 (4.80) |
| Waist circumference (cm) | 90.76 (14.01) | 91.25 (14.51) | 90.15 (13.19) | 90.66 (14.56) | 90.32 (13.48) | 90.30 (13.47) |
| Hip circumference (cm) | 103.10 (9.45) | 102.90 (9.84) | 103.40 (8.77) | 103.20 (9.33) | 103.4 (9.25) | 103.4 (9.24) |
| Currently Smoking | 153 (8.76%) | 92 (9.02%) | 47 (8.16%) | 34 (8.52%) | 38198 (7.63%) | 31573 (7.60%) |
| Exposure to smoking at home (hour/week) | 0.47 (4.07) | 0.48 (4.49) | 0.53 (3.77) | 0.42 (4.01) | 0.49 (4.44) | 0.49 (4.46) |
| Exposure to smoking out of home (hour/week) | 0.35 (2.26) | 0.37 (2.31) | 0.37 (2.41) | 0.40 (3.36) | 0.37 (2.44) | 0.37 (2.45) |
| COVID-19: Coronavirus Disease 2019, N: number, SD: standard deviation, BMI: body mass index, MVPA: self-reported moderate-to-vigorous physical activity, AMPA: acceleration vector magnitude physical activity, *: control group that excludes participants who are not in England, who died before 01/01/2020, and who have tested negative for SARS-CoV-2. |  |  |  |  |  |  |

**Table 2.** Association analysis between types physical activity and four COVID-19 related outcomes.

|  |  |  |  |  |  |  |  |  |  |  |  |  |  |  |  |  |  |  |  |
| --- | --- | --- | --- | --- | --- | --- | --- | --- | --- | --- | --- | --- | --- | --- | --- | --- | --- | --- | --- |
|  | Overall COVID-19 |  |  |  |  | Inpatients COVID-19 |  |  |  |  | Outpatients COVID-19 |  |  |  |  | COVID-19 death |  |  |  |
|  | MVPA (Subjectively measured) |  | AMPA (Objectively measured) |  |  | MVPA (Subjectively measured) |  | AMPA (Objectively measured) |  |  | MVPA (Subjectively measured) |  | AMPA (Objectively measured) |  |  | MVPA (Subjectively measured) |  | AMPA (Objectively measured) |  |
|  | OR (95%CI) | P | OR (95%CI) | P |  | OR (95%CI) | P | OR (95%CI) | P |  | OR (95%CI) | P | OR (95%CI) | P |  | OR (95%CI) | P | OR (95%CI) | P |
| Physical activity | 1.01 (0.97, 1.06) | 0.584 | **0.84 (0.73, 0.98)** | **0.021** |  | 1.05 (0.99, 1.11) | 0.104 | 0.83 (0.68, 1.00) | 0.056 |  | 0.94 (0.86, 1.02) | 0.156 | 0.92 (0.74, 1.16) | 0.495 |  | 1.03 (0.94, 1.14) | 0.493 | **0.56 (0.38, 0.83)** | **0.004** |
| Physical activity | 1.01 (0.97, 1.06) | 0.591 | **0.80 (0.69, 0.92)** | **0.003** |  | 1.05 (0.99, 1.11) | 0.105 | 0.84 (0.69, 1.02) | 0.086 |  | 0.96 (0.86, 1.02) | 0.152 | **0.74 (0.58, 0.95)** | **0.017** |  | 1.03 (0.94, 1.14) | 0.485 | 0.79 (0.53, 1.17) | 0.239 |
| Age | 1.00 (1.00, 1.01) | 0.155 | 0.97 (0.95, 0.99) | 3.09E-04 |  | 1.01 (1.01, 1.02) | 3.70E-04 | 1.00 (0.97, 1.02) | 0.754 |  | 0.96 (0.95, 0.97) | 2.81E-13 | 0.90 (0.88, 0.93) | 1.46E-11 |  | 1.13 (1.11, 1.15) | 3.47E-45 | 1.18 (1.10, 1.26) | 2.42E-06 |
| Sex | 1.34 (1.22, 1.47) | 1.09E-09 | 1.36 (1.04, 1.78) | 0.024 |  | 1.50 (1.33, 1.70) | 1.04E-10 | 1.83 (1.27, 2.62) | 0.001 |  | 1.02 (0.87, 1.21) | 0.784 | 0.84 (0.54, 1.33) | 0.468 |  | 2.00 (1.63, 2.45) | 3.41E-11 | 1.84 (0.93, 3.65) | 0.08 |
| Physical activity | 1.02 (0.97, 1.06) | 0.504 | **0.80 (0.69, 0.93)** | **0.003** |  | 1.05 (0.99, 1.11) | 0.108 | 0.85 (0.70, 1.03) | 0.096 |  | 0.95 (0.87, 1.03) | 0.233 | **0.74 (0.58, 0.95)** | **0.019** |  | 1.04 (0.94, 1.14) | 0.449 | 0.78 (0.52, 1.17) | 0.235 |
| Age | 1.00 (1.00, 1.01) | 0.156 | 0.97 (0.95, 0.99) | 2.66E-04 |  | 1.01 (1.01, 1.02) | 4.20E-04 | 1.00 (0.97, 1.02) | 0.756 |  | 0.96 (0.95, 0.97) | 2.51E-13 | 0.90 (0.87, 0.93) | 7.48E-12 |  | 1.13 (1.11, 1.15) | 3.30E-45 | 1.18 (1.10, 1.26) | 2.45E-06 |
| Sex | 1.33 (1.21, 1.47) | 2.29E-09 | 1.33 (1.02, 1.75) | 0.037 |  | 1.50 (1.32, 1.70) | 1.70E-10 | 1.80 (1.25, 2.58) | 0.002 |  | 1.01 (0.86, 1.19) | 0.882 | 0.80 (0.50, 1.28) | 0.351 |  | 1.98 (1.61, 2.43) | 6.89E-11 | 1.84 (0.93, 3.65) | 0.082 |
| Waist circumference | 1.00 (1.00, 1.01) | 0.231 | 1.00 (0.99, 1.01) | 0.557 |  | 1.00 (1.00, 1.01) | 0.05 | 1.01 (1.00, 1.03) | 0.05 |  | 1.00 (0.99, 1.01) | 0.789 | 0.99 (0.97, 1.00) | 0.097 |  | 1.00 (0.99, 1.01) | 0.78 | 1.00 (0.98, 1.03) | 0.891 |
| Hip circumference | 1.00 (0.99, 1.00) | 0.166 | 1.00 (0.99, 1.02) | 0.871 |  | 0.99 (0.99, 1.00) | 0.101 | 1.01 (0.98, 1.02) | 0.87 |  | 1.00 (0.99, 1.01) | 0.993 | 1.00 (0.97, 1.02) | 0.884 |  | 1.00 (0.99, 1.01) | 0.652 | 0.99 (0.95, 1.03) | 0.644 |
| BMI | 1.01 (1.00, 1.01) | 0.315 | 1.01 (0.98, 1.04) | 0.52 |  | 1.00 (0.99, 1.02) | 0.67 | 1.00 (0.96, 1.04) | 0.981 |  | 1.01 (0.99, 1.03) | 0.264 | 1.03 (0.98, 1.07) | 0.21 |  | 1.01 (0.99, 1.03) | 0.505 | 0.99 (0.92, 1.06) | 0.761 |
| Physical activity | 1.01 (0.95, 1.06) | 0.876 | **0.82 (0.69, 0.96)** | **0.012** |  | 1.05 (0.98, 1.11) | 0.166 | 0.85 (0.69, 1.05) | 0.129 |  | 0.93 (0.85, 1.03) | 0.155 | 0.79 (0.61, 1.03) | 0.078 |  | 1.00 (0.90, 1.11) | 0.966 | 0.74 (0.48, 1.14) | 0.172 |
| Age | 1.00 (1.00, 1.00) | 0.144 | 0.98 (0.96, 0.99) | 0.01 |  | 1.02 (1.01, 1.02) | 4.31E-04 | 1.01 (0.98, 1.03) | 0.658 |  | 0.96 (0.95, 0.97) | 2.36E-12 | 0.90 (0.87, 0.93) | 8.53E-10 |  | 1.13 (1.11, 1.16) | 4.46E-40 | 1.21 (1.12, 1.31) | 1.30E-06 |
| Sex | 1.36 (1.22, 1.50) | 5.93E-09 | 1.41 (1.05, 1.88) | 0.021 |  | 1.50 (1.31, 1.72) | 2.98E-09 | 1.94 (1.31, 2.88) | 9.22E-04 |  | 1.08 (0.91, 1.30) | 0.373 | 0.84 (0.51, 1.39) | 0.505 |  | 1.90 (1.52, 2.37) | 1.26E-08 | 1.57 (0.78, 3.18) | 0.207 |
| Waist circumference | 1.00 (1.00, 1.01) | 0.086 | 1.01 (1.00, 1.02) | 0.133 |  | 1.01 (1.00, 1.01) | 0.013 | 1.01 (1.00, 1.03) | 0.033 |  | 1.00 (0.99, 1.01) | 0.747 | 1.00 (0.98, 1.01) | 0.723 |  | 1.00 (0.99, 1.01) | 0.56 | 1.00 (0.98, 1.03) | 0.93 |
| Hip circumference | 1.00 (0.99, 1.00) | 0.127 | 1.00 (0.98, 1.01) | 0.793 |  | 0.99 (0.99, 1.00) | 0.076 | 1.00 (0.98, 1.02) | 0.775 |  | 1.00 (0.99, 1.01) | 0.962 | 0.98 (0.96, 1.01) | 0.234 |  | 1.00 (0.99, 1.01) | 0.916 | 1.00 (0.97, 1.04) | 0.821 |
| BMI | 1.00 (0.99, 1.01) | 0.449 | 1.01 (0.98, 1.04) | 0.375 |  | 1.00 (0.99, 1.02) | 0.758 | 1.01 (0.98, 1.05) | 0.456 |  | 1.01 (0.99, 1.02) | 0.486 | 1.01 (0.97, 1.06) | 0.574 |  | 1.01 (0.98, 1.03) | 0.594 | 1.01 (0.94, 1.08) | 0.844 |
| Currently smoking | 1.05 (0.93, 1.18) | 0.431 | 1.22 (0.90, 1.66) | 0.201 |  | 1.10 (0.94, 1.28) | 0.22 | 1.22 (0.81, 1.83) | 0.348 |  | 1.00 (0.81, 1.24) | 0.993 | 1.40 (0.88, 2.25) | 0.157 |  | 0.92 (0.71, 1.21) | 0.557 | 0.39 (0.16, 0.93) | 0.033 |
| Exposure to smoking at home | 1.00 (0.98, 1.01) | 0.513 | 0.95 (0.85, 1.05) | 0.291 |  | 0.99 (0.97, 1.01) | 0.368 | 0.94 (0.82, 1.09) | 0.424 |  | 1.00 (0.99, 1.02) | 0.647 | 0.96 (0.82, 1.11) | 0.559 |  | 1.00 (0.97, 1.02) | 0.879 | 0.65 (0.29, 1.45) | 0.293 |
| Exposure to smoking out of home | 1.00 (0.98, 1.02) | 0.908 | 0.91 (0.79, 1.05) | 0.192 |  | 1.00 (0.97, 1.03) | 0.962 | 0.89 (0.73, 1.09) | 0.276 |  | 1.00 (0.97, 1.04) | 0.799 | 0.80 (0.58, 1.10) | 0.169 |  | 1.01 (0.97, 1.05) | 0.671 | 1.00 (0.87, 1.15) | 0.987 |
| COVID-19: Coronavirus Disease 2019, MVPA: self-reported moderate-to-vigorous physical activity, AMPA: acceleration vector magnitude physical activity, BMI: body mass index, OR: odds ratio, CI: confidence interval |  |  |  |  |  |  |  |  |  |  |  |  |  |  |  |  |  |  |  |

**Figure 1.**
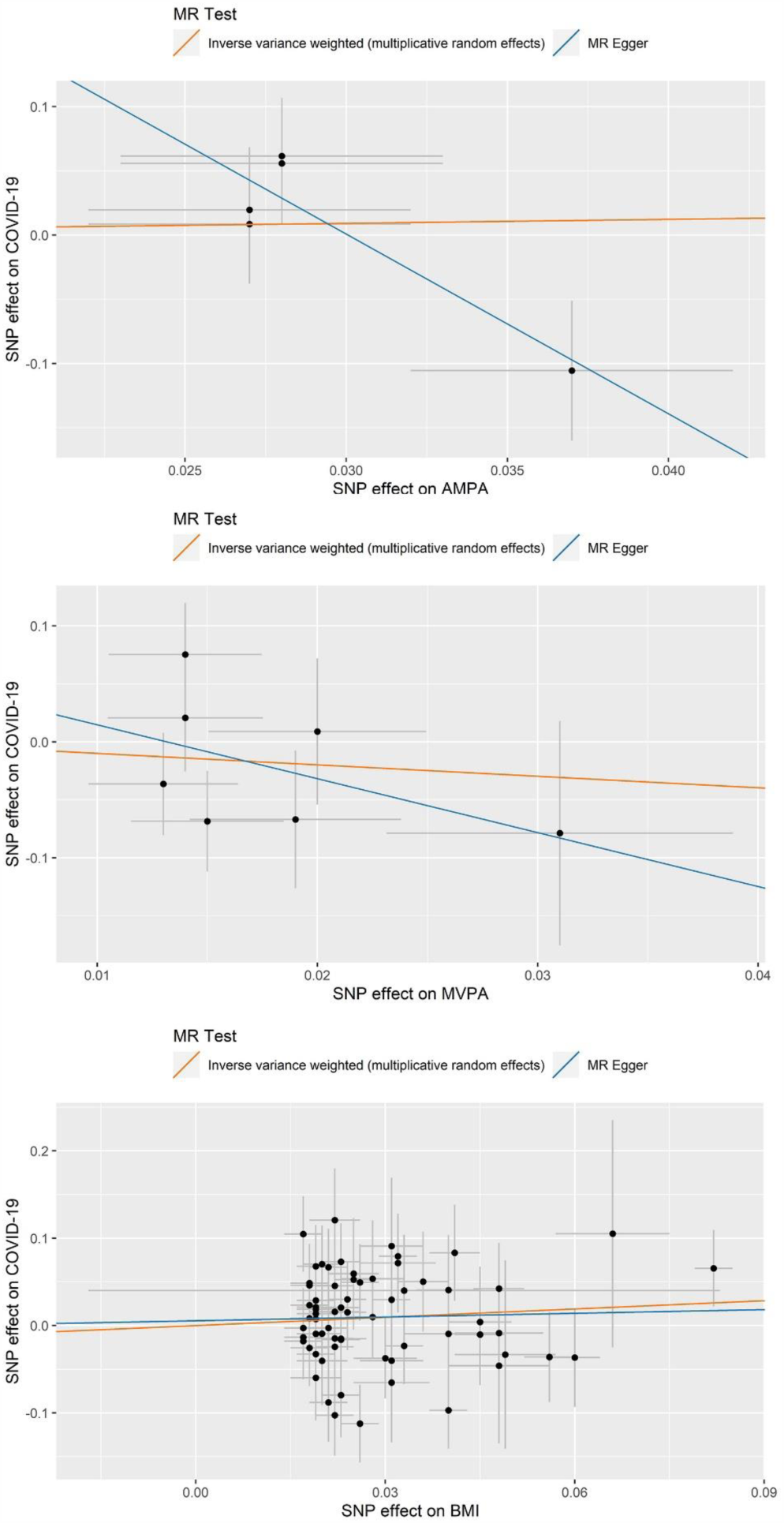
Scatter plots of results from two-sample Mendelian randomisation (MR) analysis. MVPA: moderate-to-vigorous physical activity, COVID-19: coronavirus disease 2019, AMPA: acceleration vector magnitude physical activity, BMI: body mass index.

A total of 500,758 participants (96,460 participants have AMPA data) from the UKBB were included as controls of the observational analyses. For the COVID-19 test result records, 1,596 patients were COVID-19 positive, of these, 1,020 were inpatients and 576 were outpatients. In addition, 399 participants had died of COVID-19 (with 376 deaths having COVID-19 as primary cause of death), in which 249 had SARS-CoV-2 positive results. In the multivariate logistic regression models, AMPA was associated with decreased risk of contracting COVID-19 and attending as an outpatient with a COVID-19 related health concern. The odds ratio (OR) and 95% confidence interval (95% CI) per SD increase of AMPA were 0.80 (0.69, 0.93) and 0.74 (0.58, 0.95) respectively after adjusting for age, gender, and measures of body fatness (Table 2). Further adjusting smoking status, the OR (95%CI) was 0.81 (0.69, 0.96) for association between AMPA on overall COVID-19. In the univariate model, AMPA related to a decreased risk of COVID-19 death while the association disappeared after adjusting for the other covariates. On the contrary, MVPA was not associated with any of the COVID-19 outcomes (Table 2). The observational analysis did not indicate an association between BMI and COVID-19 outcomes (Table S1). The sensitivity analyses reported similar results with the main analyses (Tables S3 & S4).

A total of 342,678 participants with genotype data were included in the MR analysis after sample quality control. Of these, 9,494 participants had COVID-19 test results. The effect estimates of the instrumental variables on COVID-19 are listed in Table S5. AMPA detected to be associated with a lower risk of inpatient COVID-19 by the MR-Egger method, however, the intercept of MR-Egger also indicated the existence of pleiotropic effects. Significant heterogeneity has not been detected by Q statistics (Figure 1 and Supplementary Table 2). MVPA and BMI were not found to be causally linked to COVID-19 outcomes in this cohort.

COVID-19: Coronavirus Disease 2019, N: number, SD: standard deviation, BMI: body mass index, MVPA: self-reported moderate-to-vigorous physical activity, AMPA: acceleration vector magnitude physical activity, *: control group that excludes participants who are not in England, who died before 01/01/2020, and who

have tested negative for SARS-CoV-2.

## Discussion

Our study found that in this UK study population higher AMPA values were inversely associated with overall COVID-19 and outpatient COVID-19 after adjusting for age, sex, measures of obesity in the multivariate model. The association between AMPA and overall COVID-19 persisted after adjusting for smoking status. No association was detected by observational analyses between MVPA or BMI and COVID-19. No causal association has been found between any type of PA or BMI and COVID-19 outcomes in the MR analysis.

Published evidence shows that PA is not only associated with better physical health through pathways such as regulating immunity^7^ but also can benefit mental health.^8^ In addition, for respiratory viral infections, PA can increase the endurance of the respiratory muscles and improve the immune response to respiratory viral antigens.^9^ One review reported that moderate-intensity exercise reduces the risk and severity of respiratory viral infections while vigorous-intensity exercise increases the risk of self-reported respiratory viral infection symptoms.^10^ Since there was limited testing capacity at the early stage of the pandemic in the UK, most (76%) of the SARS-CoV-2 testing samples came from hospitalised patients, which can be taken as a surrogate for comparably serious disease associated with COVID-19.^11^ Overall COVID-19 covered confirmed cases from inpatient and outpatient settings, and patients, who died of or with COVID-19. Inpatient COVID-19 indicated patients with serious COVID-19 while outpatient COVID-19 indicated patients with relatively milder symptoms. In our analysis, one SD increase of AMPA related to a decreased risk of overall COVID-19 and outpatient COVID-19 but not inpatient COVID-19 and COVID-19 death. The effect estimates implied that the objectively measured PA has a larger effect on relatively mild COVID-19 outcomes. Since the effect of AMPA was abrogated after adjustment for other covariates highlights the importance of the effect of age, gender, and other co-morbidities for COVID-19 death.

This is the first study to analyse the association between PA and COVID-19 quantitatively. One of the advantages of this study is that both objectively and subjectively measured PA were analysed. AMPA is widely considered as a relatively accurate measure of PA, whereas the main advantage of using MVPA is its cost-efficiency. MVPA and AMPA measures were compared in a large Norwegian study and the results indicated that people tend to report less moderate-intensity and more vigorous-intensity PA compared to AMPA regardless of gender and age.^12^ Older adults also tend to exaggerate the reports of MVPA,^12^ perhaps because fragile individuals experience stronger proprioception during exercise. In addition, self-reported questionnaires tend to record more intentional outdoor exercise while accelerometers measure all types of exercise.^13, 14^ During the pandemic these differences may intensify. Another advantage of this study was that four COVID-19 outcomes were analysed. Although more data reflecting serious diseases related to COVID-19 is available, the four outcomes can still represent different severity of COVID-19 disease.

Our study has several limitations. Firstly, participants of the UKBB tend to be healthier, leaner, and smoke less compared to the general population of the UK, especially for those who accepted the invitation to take part in the accelerometer aspect of the study. Furthermore, UKBB participants were recruited when aged 40 to 69 years old, and so may now have an even higher risk of severe COVID-19 disease. The lower obese/overweight and smoking rate and older age may result in an underestimation of the effects of obesity and smoking status. Secondly, the data for AMPA were obtained between 2013 and 2015, and older age related to a lower AMPA, which could lead to an underestimation of the true effect of AMPA on COVID-19.^12^ The data for MVPA and BMI were acquired even earlier (2006-2010), so, the historical measures may not be a good proxy for current values. Combining the discrepancy between objectively and subjectively measured PA and the time gap may explain the different results of MVPA and AMPA on COVID-19 related outcomes. Thirdly, our study did not support the enhanced effect of BMI on COVID-19, even not for the COVID-19 death outcome.^15^ This may be due to the relatively leaner UKBB population, and the time gap between BMI and COVID-19 testing. Finally, testing practices and capacity changed over time in the UK, so the earlier data (data prior to 30/05/2020 represent a limited subset of COVID-19 infections. Due to the limited number of test results, the MR analysis had low power to detect a significant association.

## Conclusion

Our results indicate a protective effect of objectively measured PA and adverse COVID-19 outcomes (outpatient COVID-19 and overall COVID-19) independent of age, sex, measures of obesity, and smoking status. Although the MR analysis did not support a causal association, that may be due to limited power. We conclude that policies to encourage and facilitate exercise at a population level during the pandemic should be considered.

## Materials and Methods

### Dataset

The UK-Biobank (UKBB) is a prospective cohort study including more than 500,000 participants aged from 40 to 69 years in the United Kingdom. In this study, participants who have not been tested positive for SARS-CoV-2 and not died of COVID-19 were taken as controls. We removed from the controls the following participants in a sensitivity analysis: i) Those who tested negative, since test results could have been false negative; ii) Participants, who were not from England, since all COVID-19 test results were provided by NHS England only; iii) Participants who died before 01/01/2020. We used both self-reported moderate-to-vigorous PA (MVPA) and acceleration vector magnitude PA (AMPA) as measures of physical activity. MVPA data were mainly acquired during 2006 to 2010 through touch screen questionnaire^16^ and AMPA data were collected from a subset of 103,687 UKBB participants wearing an accelerometer for 7 days between 2013 and 2015.^13^ For MVPA, we assigned as “NA” for individuals who selected “prefer not to answer” or “do not know” on the questions, individuals reporting being unable to walk, and individuals reporting MPA or VPA for more than 16 hours per day. We recoded those reporting >3 hours/day of MPA or VPA to 3 hours. MVPA was calculated by taking the sum of total minutes per week of MPA multiplied by four and the total number of VPA minutes per week multiplied by eight.^11^ The AMPA covered 103,687 participants in UKBB, we removed participants whose data could not be calibrated, values that were unrealistically high (average vector magnitude > 100 mg), or who had poor wear-time. BMI data were collected from 2006 to 2010 and calculated using manual measured weight and height by trained recruiters.^17^

UKBB released COVID-19 test results with the date of the specimen taken, specimen type (e.g. nasal and throat), the testing laboratory, whether the patient was an inpatient or outpatient when the sample was taken and SARS-CoV-2 testing results. Inpatient samples included inpatient infections, samples taken in emergency departments and health care related infections. From 16/03/2020 to 29/06/2020, there were 14,439 SARS-CoV-2 test results among UK Biobank participants with 1,596 participants having at least one SARS-CoV-2 positive result and 7,898 participants having one or more negative results. Of these individuals, 7,187 were inpatients and 2,307 were outpatients. We analysed four COVID-19 related outcomes: overall COVID-19, included all patients (inpatients, outpatients or deaths); inpatient COVID-19, included all inpatients with at least one positive SARS-CoV-2 testing result; outpatient COVID-19, included outpatients who tested SARS-CoV-2 positive at least once; COVID-19 death, included death caused by clinical and epidemiological diagnosed COVID-19 (both primary and contributory causes of death).

Subsequently, we performed two-sample MR analysis to test the causality between PA and BMI and COVID-19 in UKBB. The detailed description of genotyping, quality control and genetic imputation have been described previously.^18^ From a total of 488,366 participants in the UKBB with genotype data, 149,110 samples were excluded due to consent withdrawals, non-white British ethnic background, sex mismatch, sex aneuploidy, high missing rate/outlier, and kinship inference. The genetic instruments for MVPA, AMPA, and BMI were extracted from the largest available GWAS datasets.^19-21^ Seven, five, and 68 common genetic variants were extracted for MVPA, AMPA, and BMI respectively after considering linkage disequilibrium (LD) (r^2^>0.2).

### Statistical analyses

To describe the characteristics of participants, mean and standard deviation (SD) are presented for continuous covariates, and number (N) and percentage (%) are presented for categorical covariates. We performed both univariate and multivariate logistic regression analysis to test the association between two measures of PA and four COVID-19 related outcomes. In the multivariate logistic regression model, we first added age and sex as covariates, then we further added measures of obesity or overweight (waist circumference, hip circumference, and BMI), and finally, we added smoking measures (current smoking status, exposure to smoking at home and exposure to smoking out of home). The details of the covariates can be found in Bycroft et al.^17^

For the MR analysis, we tested the association between genetic instruments of the exposures and COVID-19 by performing multivariate logistic regression adjusting for age, sex, the first 10 principal components (PCs), and the assessment centre in UKBB. The causal effects and the corresponding standard errors of exposures on the outcome were calculated by using a random effect inverse-variant weighted (IVW) method.^22^ We evaluated the heterogeneity among the causal effects of each variant (Cochran’s Q statistic) and a P-value lower than 0.10^23^ was regarded as statistically significant heterogeneity. We performed MR-Egger^24^ as a sensitivity analysis to explore the potential bias introduced by horizontal pleiotropy.

The P-value threshold was set at 0.05 for all the analyses. All statistical analyses were performed on R v3.6.1.

## Data Availability

No additional data available

## Competing interests

The authors declare that they have no conflict of interest.

## Funding

This work was supported by Cancer Research UK programme grant to MD [grant number C348/A18927]; Cancer Research UK Career Development Fellowship to ET [grant number C31250/A22804]; The Darwin Trust of Edinburgh studentship to XZ.

## Acknowledgments

We would like to express our gratitude to the support from Edinburgh CRUK Cancer Research Centre as well as support from all trustees of the Darwin Trust of Edinburgh. This research has been conducted using the UK Biobank Resource under Application Number 10775.

---

**Table S1.**
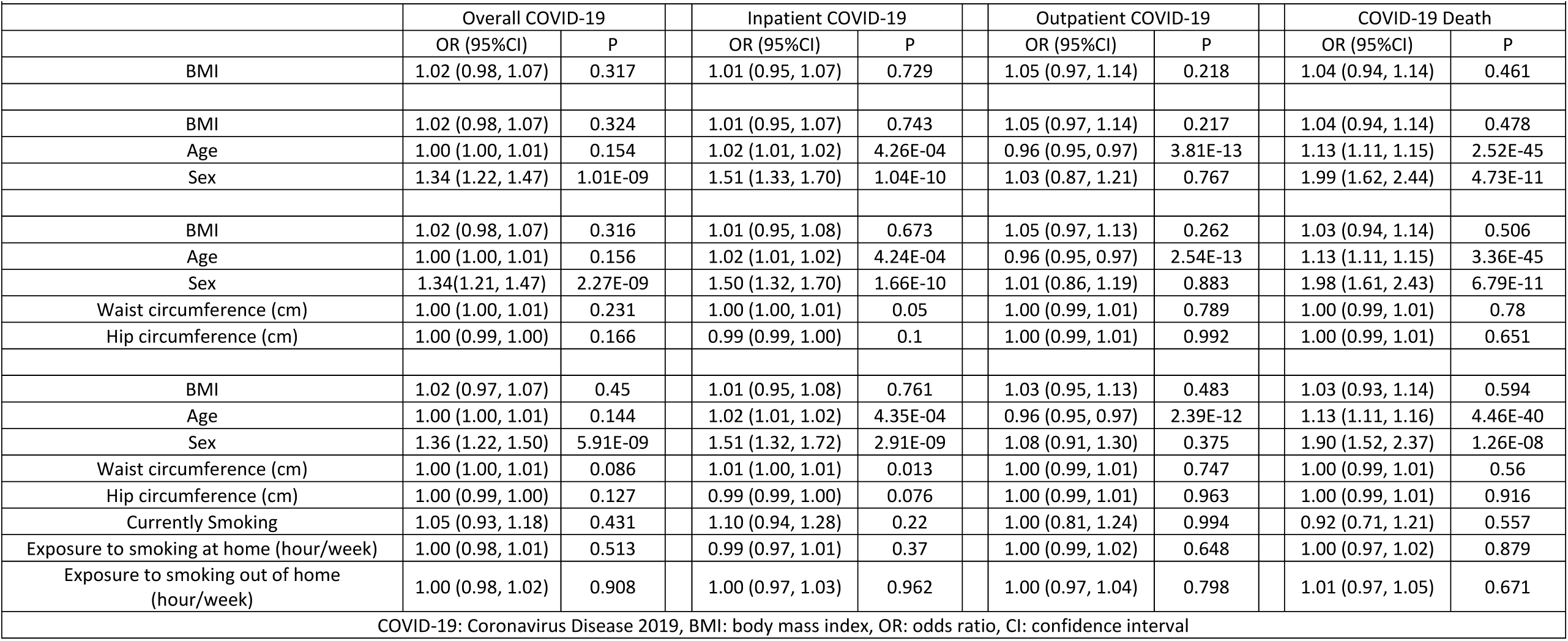
Association analysis between obesity measures and four COVID-19 related outcomes.

**Table S2.**
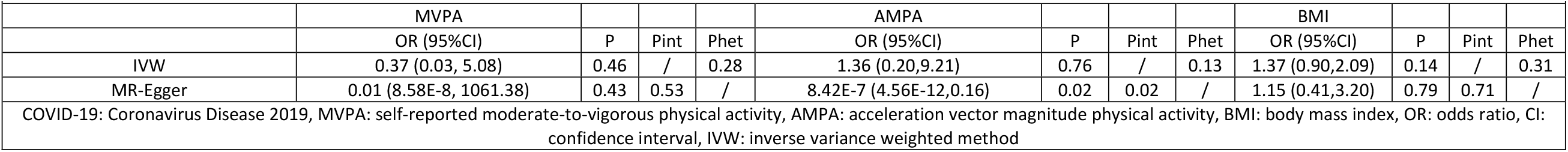
Results of two-sample Mendelian randomisation studies between physical activity exposures or BMI and COVID-19 outcomes.

**Table S3.**
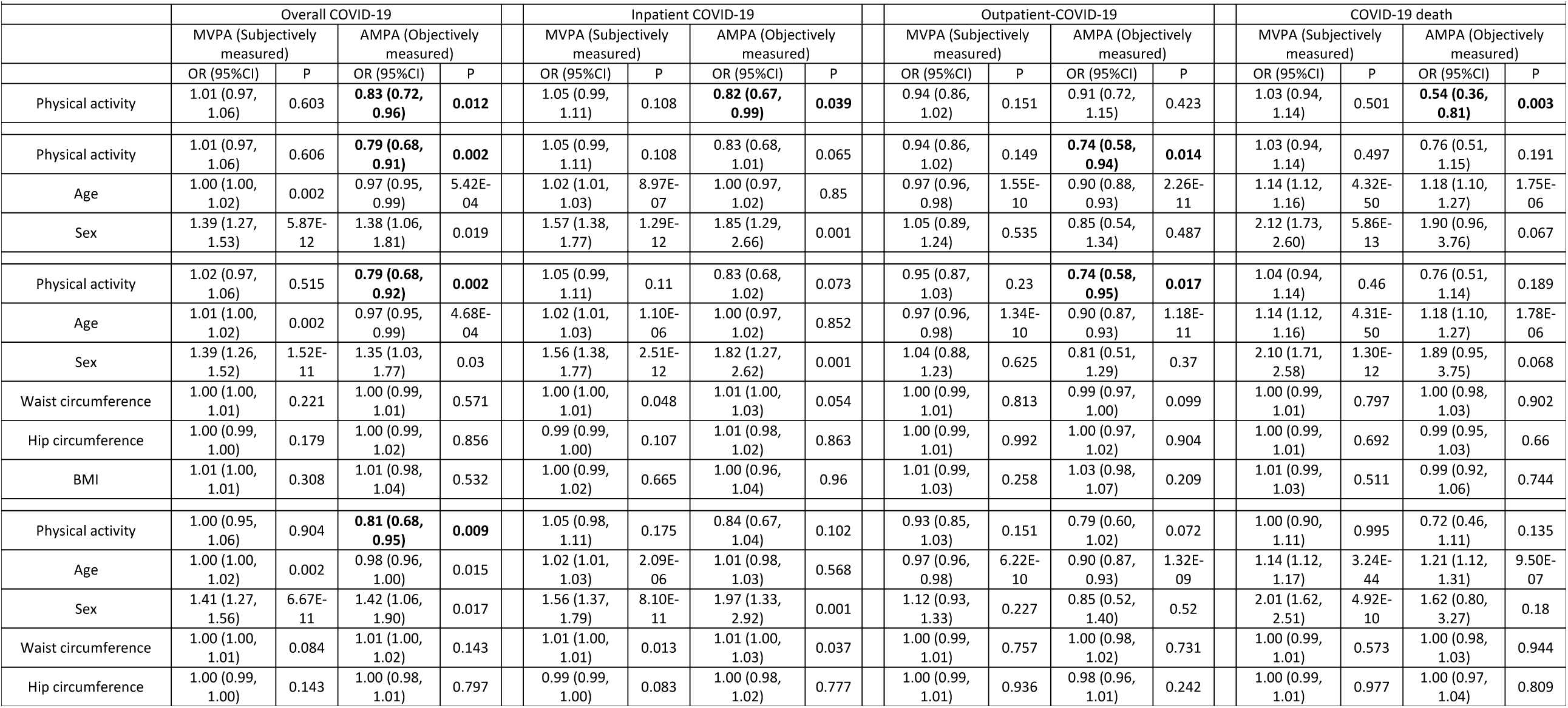
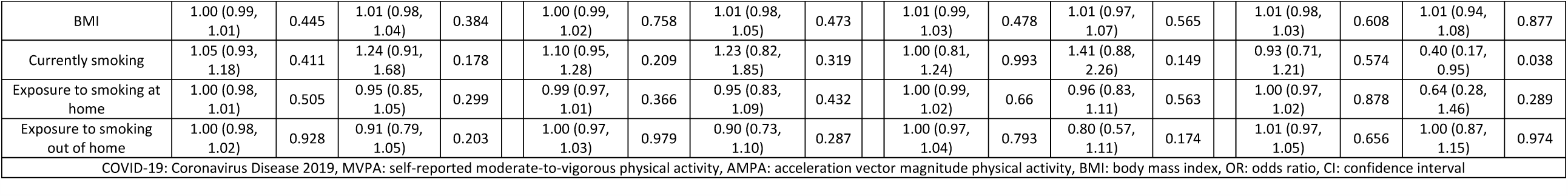
Sensitivity analysis for the association between physical activity measures and four COVID-19 related outcomes after excluding controls not from England, that died before 01/01/2020, and participants that tested negative for SARS-CoV-2.

**Table S4.**
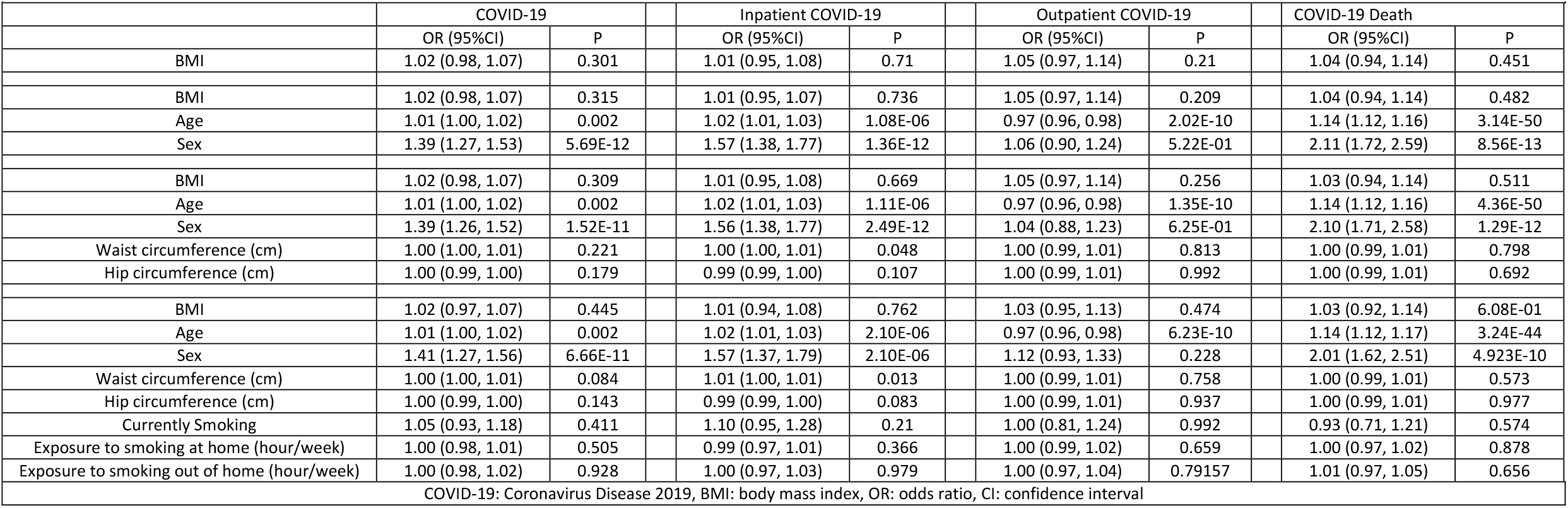
Association analysis between BMI and four COVID-19 related outcomes after excluding controls not from England, that died before 01/01/2020, and participants that tested negative for SARS-CoV-2.

**Table S5.** Effect estimates of each instrumental variable of each exposure (MVPA, AMPA, BMI) on the overall COVID-19 outcome.

|  |  |  |  |  |  |  |  |  |  |  |  |
| --- | --- | --- | --- | --- | --- | --- | --- | --- | --- | --- | --- |
| SNP | Exposure | BP | CHR | EA | EAF | Beta.PA | SE.PA | Beta.BMI | SE.BMI | Beta.COVID | SE.COVID |
| rs149943 | MVPA | 28002388 | 6 | G/A | 0.85 | 0.019 | 0.005 | / | / | -0.067 | 0.059 |
| rs2035562 | MVPA | 85056521 | 3 | A/G | 0.33 | -0.014 | 0.004 | / | / | -0.021 | 0.046 |
| rs2854277 | MVPA | 32628084 | 6 | C/T | 0.92 | 0.031 | 0.008 | / | / | -0.079 | 0.097 |
| rs2988004 | MVPA | 37044388 | 9 | T/G | 0.56 | -0.013 | 0.003 | / | / | 0.036 | 0.044 |
| rs3094622 | MVPA | 30327952 | 6 | A/G | 0.86 | 0.02 | 0.005 | / | / | 0.009 | 0.063 |
| rs7791992 | MVPA | 50237784 | 7 | C/A | 0.41 | -0.014 | 0.003 | / | / | -0.075 | 0.044 |
| rs7804463 | MVPA | 133447651 | 7 | T/C | 0.53 | 0.015 | 0.003 | / | / | -0.068 | 0.043 |
| rs59499656 | AMPA | 40768309 | 18 | A/T | 0.655 | -0.028 | 0.005 | / | / | -0.062 | 0.045 |
| rs6895232 | AMPA | 152039421 | 5 | T/A | 0.663 | 0.027 | 0.005 | / | / | 0.009 | 0.046 |
| rs564819152 | AMPA | 21820650 | 10 | A/G | 0.679 | 0.028 | 0.005 | / | / | 0.056 | 0.047 |
| rs55657917 | AMPA | 44326864 | 17 | A/G | 0.77 | -0.037 | 0.005 | / | / | 0.106 | 0.054 |
| rs6775319 | AMPA | 18758501 | 3 | A/T | 0.271 | 0.027 | 0.005 | / | / | 0.02 | 0.049 |
| rs1000940 | BMI | 5283252 | 17 | G/A | 0.32 | / | / | 0.019 | 0.003 | -0.009 | 0.047 |
| rs10938397 | BMI | 45182527 | 4 | G/A | 0.434 | / | / | 0.04 | 0.003 | -0.097 | 0.044 |
| rs10968576 | BMI | 28414339 | 9 | G/A | 0.32 | / | / | 0.025 | 0.003 | 0.053 | 0.046 |
| rs11126666 | BMI | 26928811 | 2 | A/G | 0.283 | / | / | 0.021 | 0.003 | -0.003 | 0.05 |
| rs11165643 | BMI | 96924097 | 1 | T/C | 0.583 | / | / | 0.022 | 0.003 | -0.015 | 0.044 |
| rs11191560 | BMI | 104869038 | 10 | C/T | 0.089 | / | / | 0.031 | 0.005 | 0.091 | 0.078 |
| rs1167827 | BMI | 75163169 | 7 | G/A | 0.553 | / | / | 0.02 | 0.003 | -0.009 | 0.044 |
| rs11847697 | BMI | 30515112 | 14 | T/C | 0.042 | / | / | 0.049 | 0.008 | -0.033 | 0.108 |
| rs12286929 | BMI | 115022404 | 11 | G/A | 0.523 | / | / | 0.022 | 0.003 | 0.045 | 0.043 |
| rs12401738 | BMI | 78446761 | 1 | A/G | 0.352 | / | / | 0.021 | 0.003 | 0.067 | 0.044 |
| rs12429545 | BMI | 54102206 | 13 | A/G | 0.133 | / | / | 0.033 | 0.05 | 0.04 | 0.064 |
| rs13021737 | BMI | 632348 | 2 | G/A | 0.828 | / | / | 0.06 | 0.004 | -0.037 | 0.057 |
| rs13107325 | BMI | 103188709 | 4 | T/C | 0.072 | / | / | 0.048 | 0.007 | -0.009 | 0.083 |
| rs1516725 | BMI | 185824004 | 3 | C/T | 0.872 | / | / | 0.045 | 0.005 | 0.004 | 0.063 |
| rs1528435 | BMI | 181550962 | 2 | T/C | 0.631 | / | / | 0.018 | 0.003 | 0.049 | 0.045 |
| rs1558902 | BMI | 53803574 | 16 | A/T | 0.415 | / | / | 0.082 | 0.003 | 0.065 | 0.044 |
| rs16851483 | BMI | 141275436 | 3 | T/G | 0.066 | / | / | 0.048 | 0.008 | -0.046 | 0.089 |
| rs17001654 | BMI | 77129568 | 4 | G/C | 0.153 | / | / | 0.031 | 0.005 | -0.04 | 0.063 |
| rs17024393 | BMI | 110154688 | 1 | C/T | 0.04 | / | / | 0.066 | 0.009 | 0.105 | 0.13 |
| rs17094222 | BMI | 102395440 | 10 | C/T | 0.211 | / | / | 0.025 | 0.004 | 0.059 | 0.052 |
| rs2033529 | BMI | 40348653 | 6 | G/A | 0.293 | / | / | 0.019 | 0.003 | 0.068 | 0.047 |
| rs2033732 | BMI | 85079709 | 8 | C/T | 0.747 | / | / | 0.019 | 0.004 | -0.06 | 0.049 |
| rs205262 | BMI | 34563164 | 6 | G/A | 0.273 | / | / | 0.022 | 0.004 | 0.016 | 0.049 |
| rs2121279 | BMI | 143043285 | 2 | T/C | 0.152 | / | / | 0.025 | 0.004 | 0.059 | 0.064 |
| rs2207139 | BMI | 50845490 | 6 | G/A | 0.177 | / | / | 0.045 | 0.004 | -0.01 | 0.058 |
| rs29941 | BMI | 34309532 | 19 | G/A | 0.669 | / | / | 0.018 | 0.003 | 0.008 | 0.046 |
| rs3101336 | BMI | 72751185 | 1 | C/T | 0.613 | / | / | 0.033 | 0.003 | -0.023 | 0.044 |
| rs3810291 | BMI | 47569003 | 19 | A/G | 0.666 | / | / | 0.028 | 0.004 | 0.01 | 0.046 |
| rs3817334 | BMI | 47650993 | 11 | T/C | 0.407 | / | / | 0.026 | 0.003 | 0.049 | 0.044 |
| rs3849570 | BMI | 81792112 | 3 | A/C | 0.359 | / | / | 0.019 | 0.003 | 0.013 | 0.045 |
| rs3888190 | BMI | 28889486 | 16 | A/C | 0.403 | / | / | 0.031 | 0.003 | 0.03 | 0.044 |
| rs4256980 | BMI | 8673939 | 11 | G/C | 0.646 | / | / | 0.021 | 0.003 | -0.088 | 0.045 |
| rs543874 | BMI | 177889480 | 1 | G/A | 0.193 | / | / | 0.048 | 0.004 | 0.042 | 0.053 |
| rs6567160 | BMI | 57829135 | 18 | C/T | 0.236 | / | / | 0.056 | 0.004 | -0.036 | 0.052 |
| rs6804842 | BMI | 25106437 | 3 | G/A | 0.575 | / | / | 0.019 | 0.003 | -0.033 | 0.044 |
| rs7138803 | BMI | 50247468 | 12 | A/G | 0.384 | / | / | 0.032 | 0.003 | 0.079 | 0.044 |
| rs7141420 | BMI | 79899454 | 14 | T/C | 0.527 | / | / | 0.024 | 0.003 | 0.015 | 0.044 |
| rs758747 | BMI | 3627358 | 16 | T/C | 0.265 | / | / | 0.023 | 0.004 | -0.015 | 0.049 |
| rs7899106 | BMI | 87410904 | 10 | G/A | 0.052 | / | / | 0.04 | 0.007 | -0.01 | 0.099 |
| rs9400239 | BMI | 108977663 | 6 | C/T | 0.688 | / | / | 0.019 | 0.003 | 0.019 | 0.048 |
| rs10132280 | BMI | 25928179 | 14 | C/A | 0.682 | / | / | 0.023 | 0.003 | -0.016 | 0.047 |
| rs1016287 | BMI | 59305625 | 2 | T/C | 0.287 | / | / | 0.023 | 0.003 | -0.08 | 0.048 |
| rs10733682 | BMI | 129460914 | 9 | A/G | 0.478 | / | / | 0.017 | 0.003 | -0.018 | 0.044 |
| rs11030104 | BMI | 27684517 | 11 | A/G | 0.792 | / | / | 0.041 | 0.004 | 0.083 | 0.055 |
| rs11057405 | BMI | 122781897 | 12 | G/A | 0.901 | / | / | 0.031 | 0.006 | -0.065 | 0.069 |
| rs11688816 | BMI | 63053048 | 2 | G/A | 0.525 | / | / | 0.017 | 0.003 | -0.014 | 0.044 |
| rs1218793 | BMI | 28007830 | 13 | T/C | 0.203 | / | / | 0.03 | 0.005 | -0.038 | 0.046 |
| rs12446632 | BMI | 19935389 | 16 | G/A | 0.865 | / | / | 0.04 | 0.005 | 0.041 | 0.063 |
| rs12566985 | BMI | 75002193 | 1 | G/A | 0.446 | / | / | 0.024 | 0.003 | 0.03 | 0.044 |
| rs12940622 | BMI | 78615571 | 17 | G/A | 0.575 | / | / | 0.018 | 0.003 | -0.026 | 0.043 |
| rs13191362 | BMI | 163033350 | 6 | A/G | 0.879 | / | / | 0.028 | 0.005 | 0.054 | 0.067 |
| rs17405819 | BMI | 76806584 | 8 | T/C | 0.7 | / | / | 0.022 | 0.003 | -0.103 | 0.046 |
| rs17724992 | BMI | 18454825 | 19 | A/G | 0.746 | / | / | 0.019 | 0.004 | 0.007 | 0.049 |
| rs1808579 | BMI | 21104888 | 18 | C/T | 0.534 | / | / | 0.017 | 0.003 | 0.105 | 0.043 |
| rs1928295 | BMI | 120378483 | 9 | T/C | 0.548 | / | / | 0.019 | 0.003 | 0.029 | 0.044 |
| rs2112347 | BMI | 75015242 | 5 | T/G | 0.629 | / | / | 0.026 | 0.003 | -0.112 | 0.045 |
| rs2176598 | BMI | 43864278 | 11 | T/C | 0.251 | / | / | 0.02 | 0.004 | -0.041 | 0.051 |
| rs2245368 | BMI | 76608143 | 7 | C/T | 0.18 | / | / | 0.032 | 0.006 | 0.071 | 0.056 |
| rs2287019 | BMI | 46202172 | 19 | C/T | 0.804 | / | / | 0.036 | 0.004 | 0.05 | 0.057 |
| rs2365389 | BMI | 61236462 | 3 | C/T | 0.582 | / | / | 0.02 | 0.003 | 0.07 | 0.044 |
| rs3736485 | BMI | 51748610 | 15 | A/G | 0.454 | / | / | 0.018 | 0.003 | 0.023 | 0.044 |
| rs4740619 | BMI | 15634326 | 9 | T/C | 0.542 | / | / | 0.018 | 0.003 | 0.046 | 0.044 |
| rs6477694 | BMI | 111932342 | 9 | C/T | 0.365 | / | / | 0.017 | 0.003 | -0.003 | 0.045 |
| rs657452 | BMI | 49589847 | 1 | A/G | 0.394 | / | / | 0.023 | 0.003 | 0.073 | 0.044 |
| rs7243357 | BMI | 56883319 | 18 | T/G | 0.812 | / | / | 0.022 | 0.004 | 0.121 | 0.059 |
| rs7599312 | BMI | 213413231 | 2 | G/A | 0.724 | / | / | 0.022 | 0.003 | -0.024 | 0.049 |
| rs7903146 | BMI | 114758349 | 10 | C/T | 0.713 | / | / | 0.023 | 0.003 | 0.021 | 0.048 |
| rs9925964 | BMI | 31129895 | 16 | A/G | 0.62 | / | / | 0.019 | 0.003 | 0.021 | 0.045 |
| SNP: single-nucleotide polymorphism, EA: effect allele, EAF: effect allele frequency, PA: physical activity, SE: standard error, COVID: Coronavirus Disease 2019, MVPA: self-reported moderate-to-vigorous physical activity, AMPA: acceleration vector magnitude physical activity, BMI: body mass index |  |  |  |  |  |  |  |  |  |  |  |

## Notes

### Competing Interest Statement

The authors have declared no competing interest.

### Author Declarations

The research activities of UK Biobank were approved by the North West Multi-centre Research Ethics Committee (11/NW/0382) in relation to the process of participant invitation, assessment and follow-up procedures.

## Reference

1. World Health Organization. Coronavirus disease (COVID-2019) situation reports 2020 [Available from: https://www.who.int/docs/default-source/coronaviruse/situation-reports/20200729-covid-19-sitrep-191.pdf?sfvrsn=2c327e9e_2. [29/07] [2020]

2. Lesser IA, Nienhuis CP. The Impact of COVID-19 on Physical Activity Behavior and Well-Being of Canadians. Int J Environ Res Public Health. 2020;17(11). https://www.ncbi.nlm.nih.gov/pubmed/32486380.

3. Rezende LFM, Sa TH, Markozannes G, et al. Physical activity and cancer: an umbrella review of the literature including 22 major anatomical sites and 770 000 cancer cases. Br J Sports Med. 2018;52(13):826–33. https://www.ncbi.nlm.nih.gov/pubmed/29146752.

4. Vasankari V, Husu P, Vaha-Ypya H, et al. Association of objectively measured sedentary behaviour and physical activity with cardiovascular disease risk. Eur J Prev Cardiol. 2017;24(12):1311–8. https://www.ncbi.nlm.nih.gov/pubmed/28530126.

5. Sellami M, Gasmi M, Denham J, et al. Effects of Acute and Chronic Exercise on Immunological Parameters in the Elderly Aged: Can Physical Activity Counteract the Effects of Aging? Front Immunol. 2018;9:2187. https://www.ncbi.nlm.nih.gov/pubmed/30364079.

6. Carter SJ, Baranauskas MN, Fly AD. Considerations for Obesity, Vitamin D, and Physical Activity Amid the COVID-19 Pandemic. Obesity (Silver Spring, Md). 2020;28(7):1176–7. https://www.ncbi.nlm.nih.gov/pubmed/32299148.

7. Simpson RJ, Kunz H, Agha N, et al. Exercise and the Regulation of Immune Functions. Prog Mol Biol Transl Sci. 2015;135:355–80. https://www.ncbi.nlm.nih.gov/pubmed/26477922.

8. White RL, Babic MJ, Parker PD, et al. Domain-Specific Physical Activity and Mental Health: A Meta-analysis. Am J Prev Med. 2017;52(5):653–66. https://www.ncbi.nlm.nih.gov/pubmed/28153647.

9. Jakobsson J, Malm C, Furberg M, et al. Physical Activity During the Coronavirus (COVID-19) Pandemic: Prevention of a Decline in Metabolic and Immunological Functions. Front sports act living. 2020;2(57). https://www.frontiersin.org/article/10.3389/fspor.2020.00057.

10. Martin SA, Pence BD, Woods JA. Exercise and respiratory tract viral infections. Exerc Sport Sci Rev. 2009;37(4):157–64. https://www.ncbi.nlm.nih.gov/pubmed/19955864.

11. Biobank U. Records of COVID-19 test results 2020 [Available from: http://biobank.ndph.ox.ac.uk/showcase/field.cgi?id=40100. [25/06] [2020]

12. Dyrstad SM, Hansen BH, Holme IM, et al. Comparison of self-reported versus accelerometer-measured physical activity. Med Sci Sports Exerc. 2014;46(1):99–106. https://www.ncbi.nlm.nih.gov/pubmed/23793232.

13. Doherty A, Jackson D, Hammerla N, et al. Large Scale Population Assessment of Physical Activity Using Wrist Worn Accelerometers: The UK Biobank Study. PLoS One. 2017;12(2):e0169649. https://www.ncbi.nlm.nih.gov/pubmed/28146576.

14. Craig CL, Marshall AL, Sjostrom M, et al. International physical activity questionnaire: 12-country reliability and validity. Med Sci Sports Exerc. 2003;35(8):1381–95. https://www.ncbi.nlm.nih.gov/pubmed/12900694.

15. Palaiodimos L, Kokkinidis DG, Li W, et al. Severe obesity, increasing age and male sex are independently associated with worse in-hospital outcomes, and higher in-hospital mortality, in a cohort of patients with COVID-19 in the Bronx, New York. Metabolism: clinical and experimental. 2020;108:154262. https://www.ncbi.nlm.nih.gov/pubmed/32422233.

16. Sudlow C, Gallacher J, Allen N, et al. UK biobank: an open access resource for identifying the causes of a wide range of complex diseases of middle and old age. PLoS Med. 2015;12(3):e1001779. https://www.ncbi.nlm.nih.gov/pubmed/25826379.

17. Bycroft C, Freeman C, Petkova D, et al. The UK Biobank resource with deep phenotyping and genomic data. Nature. 2018;562(7726):203–9. https://www.ncbi.nlm.nih.gov/pubmed/30305743.

18. Meng X, Li X, Timofeeva MN, et al. Phenome-wide Mendelian-randomization study of genetically determined vitamin D on multiple health outcomes using the UK Biobank study. Int J Epidemiol. 2019;48(5):1425–34. https://www.ncbi.nlm.nih.gov/pubmed/31518429.

19. Klimentidis YC, Raichlen DA, Bea J, et al. Genome-wide association study of habitual physical activity in over 377,000 UK Biobank participants identifies multiple variants including CADM2 and APOE. Int J Obes (Lond). 2018;42(6):1161–76. https://www.ncbi.nlm.nih.gov/pubmed/29899525.

20. Doherty A, Smith-Byrne K, Ferreira T, et al. GWAS identifies 14 loci for device-measured physical activity and sleep duration. Nat Commun. 2018;9(1):5257. https://www.ncbi.nlm.nih.gov/pubmed/30531941.

21. Locke AE, Kahali B, Berndt SI, et al. Genetic studies of body mass index yield new insights for obesity biology. Nature. 2015;518(7538):197–206. https://www.ncbi.nlm.nih.gov/pubmed/25673413.

22. Burgess S, Scott RA, Timpson NJ, et al. Using published data in Mendelian randomization: a blueprint for efficient identification of causal risk factors. Eur J Epidemiol. 2015;30(7):543–52. https://www.ncbi.nlm.nih.gov/pubmed/25773750.

23. Higgins J, Green S. Chapter 9: Analysing data and undertaking meta-analyses. In: Jonathan J, Higgins J, Douglas G, editors. Cochrane Handbook for Systematic Reviews of Interventions Version 5.1.0 [updated March 2011]. The Cochrane Collaboration, 2011 Mar. p. 28.

24. Bowden J, Davey Smith G, Burgess S. Mendelian randomization with invalid instruments: effect estimation and bias detection through Egger regression. Int J Epidemiol. 2015;44(2):512–25. https://www.ncbi.nlm.nih.gov/pubmed/26050253.

